# Obese patient imaging: improved dose efficiency with photon-counting CT

**DOI:** 10.64898/2025.12.22.25342398

**Authors:** Leening P. Liu, Kai Mei, Shobhit Sharma, Steven Ross, Sandra S. Halliburton, Richard Thompson, Naruomi Akino, Ali H. Dhanaliwala, Leonid Roshkovan, Harold I. Litt, Peter B. Noël

## Abstract

**Objective:** To evaluate the dose efficiency of cadmium-zinc-telluride (CZT) based photon-counting CT (PCCT) compared to energy-integrating detector CT (EID-CT) across phantom sizes.

**Methods:** A patient-specific 3D-printed pancreas phantom and a phantom with tissue mimicking inserts were placed in extension rings corresponding to the 50^th^, 75^th^, 85^th^, and 95^th^ percentile adult waist circumferences. Phantoms were scanned on both PCCT and EID-CT with CTDI_vol_ ranging from 0.5 to 19.4 mGy. Noise was measured in both phantoms to evaluate dose efficiency. Non-Poisson noise at low doses (<2 mGy) was quantified using root mean square error from linear fits of the noise-dose relationship. Potential dose reduction was then assessed by matching noise levels between scanners across phantom sizes.

**Results:** PCCT demonstrated reduced noise compared to EID-CT across all phantom sizes and doses with average noise reductions of 22%, 23%, 25%, and 28% for the 50^th^, 75^th^, 85^th^, and 95^th^ percentile phantoms, respectively. Noise reduction intensified at lower doses and larger phantom sizes, reaching 88 HU at 1 mGy for the 95^th^ percentile phantom. Non-Poisson noise decreased significantly with PCCT compared to EID-CT for all phantom sizes (p < 0.013). At matched noise levels, PCCT enabled dose reductions of 33% and 44% for the 50^th^ and 95^th^ percentile phantoms, respectively.

**Conclusions:** PCCT exhibited superior dose efficiency compared to EID-CT across a range of phantom sizes. The enhanced dose efficiency enables both noise reduction and potential dose reduction for the imaging of obese patients and low-dose imaging applications.

**Key Points:** *Question:* Photon-counting CT (PCCT) enables improved quantum detection and eliminates electronic noise, but its benefits have not been evaluated for obese patient sizes.

*Findings:* Compared to EID-CT, PCCT enhanced dose efficiency with noise reduction and potential dose reduction across all phantom sizes and doses.

*Clinical Relevance:* The improved dose efficiency of PCCT facilitates noise reduction that enables diagnostic image quality, and thereby the diagnostic accuracy, for obese patients.

## Introduction

Since 1990, worldwide adult obesity has more than doubled while adolescent obesity has quadrupled^1^. The rise in obesity prevalence has led to a growing population of patients with increased health risks, including heart disease and certain cancers^2–4^. Thus, more patients will likely require CT imaging for diagnosis and disease monitoring. Of the body regions, the abdomen/pelvis is among the most demanding to image: similarly attenuating abdominal tissues with little inherent contrast necessitate low noise levels to detect lesions^5^. Standard practice for lowering noise includes increasing radiation dose with patient size to account for increased attenuation^6^. However, in obese patients, even the highest doses achievable with available scanner technology have historically struggled to yield optimal image quality for abdominal imaging^7^. Over the years, both hardware^8^ and software^9,10^ solutions for minimizing image noise have been investigated to achieve optimal image quality and diagnostic accuracy at lower doses. With further development of these technological solutions, the imaging of obese patients may be improved by ensuring diagnostic image quality even at a reduced dose.

Of the hardware solutions for noise reduction, photon-counting CT (PCCT) constitutes a significant paradigm shift in CT. Compared to traditional energy-integrating detector CT (EID-CT), PCCT employs photon-counting detectors that measure the energy of individual photons, signifying superior quantum detection^11–13^. The ability to measure photon energies further facilitates the removal of background electronic noise^11,13,14^. With EID-CT, this non-Poisson distributed electronic noise significantly affects image quality and impairs quantitative accuracy when photon counts are low, i.e. low radiation doses and increased attenuation. This effect hampers the diagnostic accuracy for obese patients and limits the widespread adoption of low-dose imaging. However, with PCCT, low energy photons associated with electronic noise can be eliminated by setting an energy threshold around 25 keV^15^. Both of these detector capabilities contribute to the improved dose efficiency of PCCT. In phantoms, several studies have demonstrated the resulting overall noise reduction, decreased electronic noise, and improved detectability of simulated lesions with PCCT compared to EID-CT^14,16–19^. These advantages translated to dose reduction in patient studies where image quality was maintained or improved, with varying degrees of dose reduction depending on the body region and CT systems^20–22^.

Across both phantom and patient studies, dose efficiency of PCCT has been assessed primarily for the average sized patient^14,16–22^, with, to our knowledge, no dedicated study using obese patients. This study aims to evaluate the dose efficiency of a cadmium zinc telluride (CZT)-based PCCT prototype (Canon Medical Systems Corporation) using lifelike 3D-printed phantoms with sizes representative of adult waist circumferences up to the 95^th^ percentile of the United States population. Our results demonstrate enhanced dose efficiency of PCCT, supporting diagnostic image quality and, consequently, improved diagnostic accuracy in obese patients.

## Methods

### Phantoms

A patient-specific *PixelPrint* pancreatic adenocarcinoma phantom was 3D printed to mimic the Hounsfield units and image texture observed in a patient scan that included both non-tumor and tumor regions^23,24^. Original patient data were selected by a board-certified abdominal radiologist and acquired on an ultra–high-resolution EID-CT system (Aquilion Precision CT, Canon Medical Systems Corporation). The pancreas phantom was then placed in a 3D-printed extension ring with an outer diameter of 20 cm (Figure 1). In addition, a 20 cm diameter phantom with tissue-mimicking and material-specific inserts (Multi-energy CT phantom, Gammex) was additionally placed in the extension ring. The phantom consisted of the following inserts: adipose, brain, brain 70, brain 100, blood + iodine 2 mg/mL, blood + iodine 4 mg/mL, calcium 50 mg/mL, iodine 2 mg/mL, iodine 5 mg/mL, and iodine 10 mg/mL. To mimic an adult waist circumference of the 50^th^ and 75^th^ percentiles of the United States population^25^, both phantoms were placed within a 32 x 32 cm 3D-printed extension ring and a 30 x 40 cm extension ring (Multi-energy CT phantom, Gammex), respectively (Figure 1AB). To mimic an adult waist circumference of the 85^th^ and 95^th^ percentiles of the United States population^25^, the 75^th^ percentile phantom was placed within 3D-printed elliptical extension rings measuring 32 x 42 and 37 x 47 cm, respectively (Figure 1CD). These phantom sizes corresponded to water equivalent diameters of 32, 34, 37, and 41 cm, respectively.

**Figure 1.**
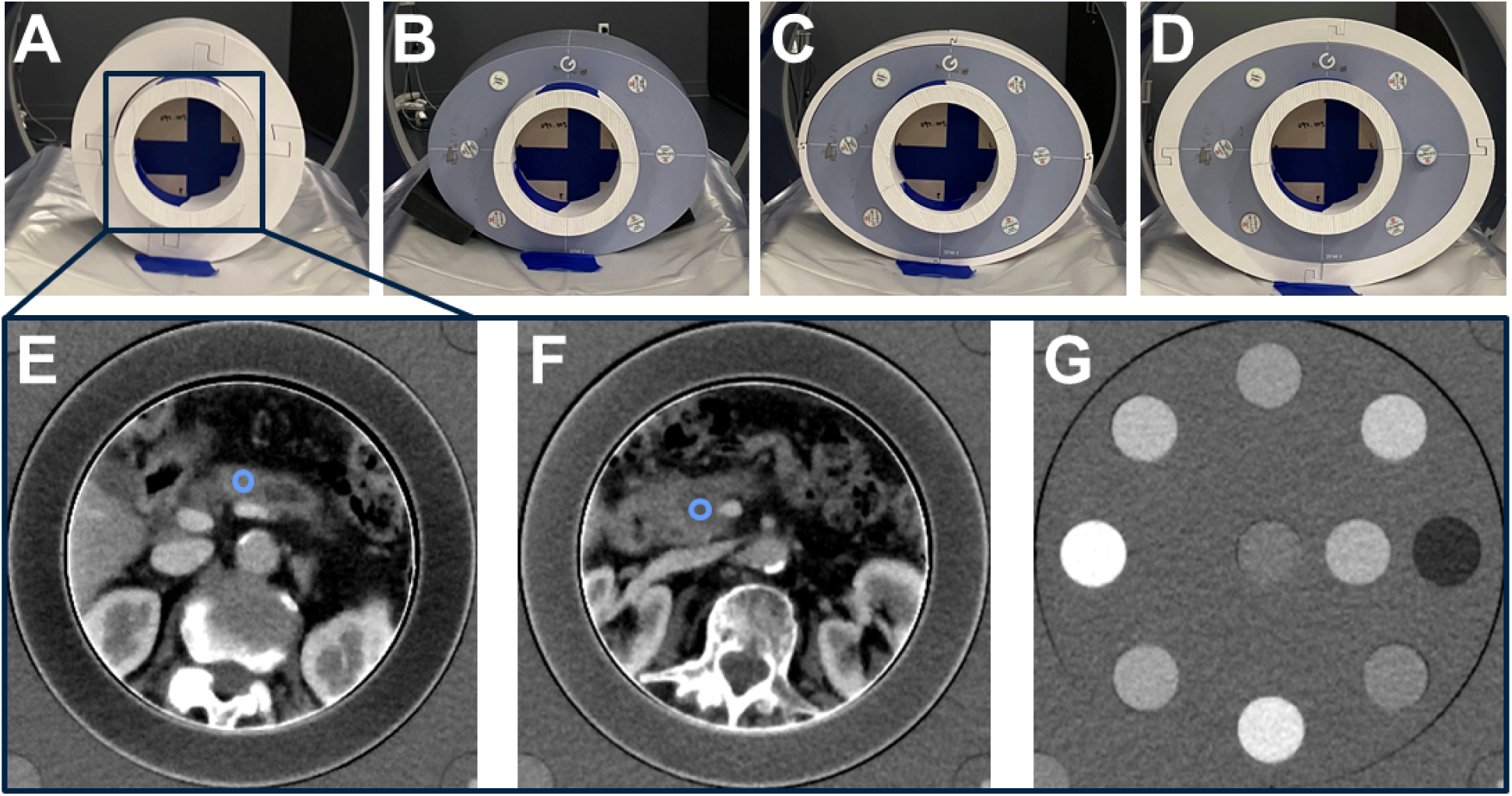
Experimental setup. A 3D-printed anthropomorphic pancreas phantom and multienergy CT phantom were scanned within extension rings to mimic the waist circumference of the 50^th^ (A), 75^th^ (B), 85^th^ (C), and 95^th^ (D) percentile of adults in the United States. The phantom was analyzed in both non-tumor region (E), tumor (F), and different inserts (G). WL/WW: 40/350 HU.

**Figure 2.**
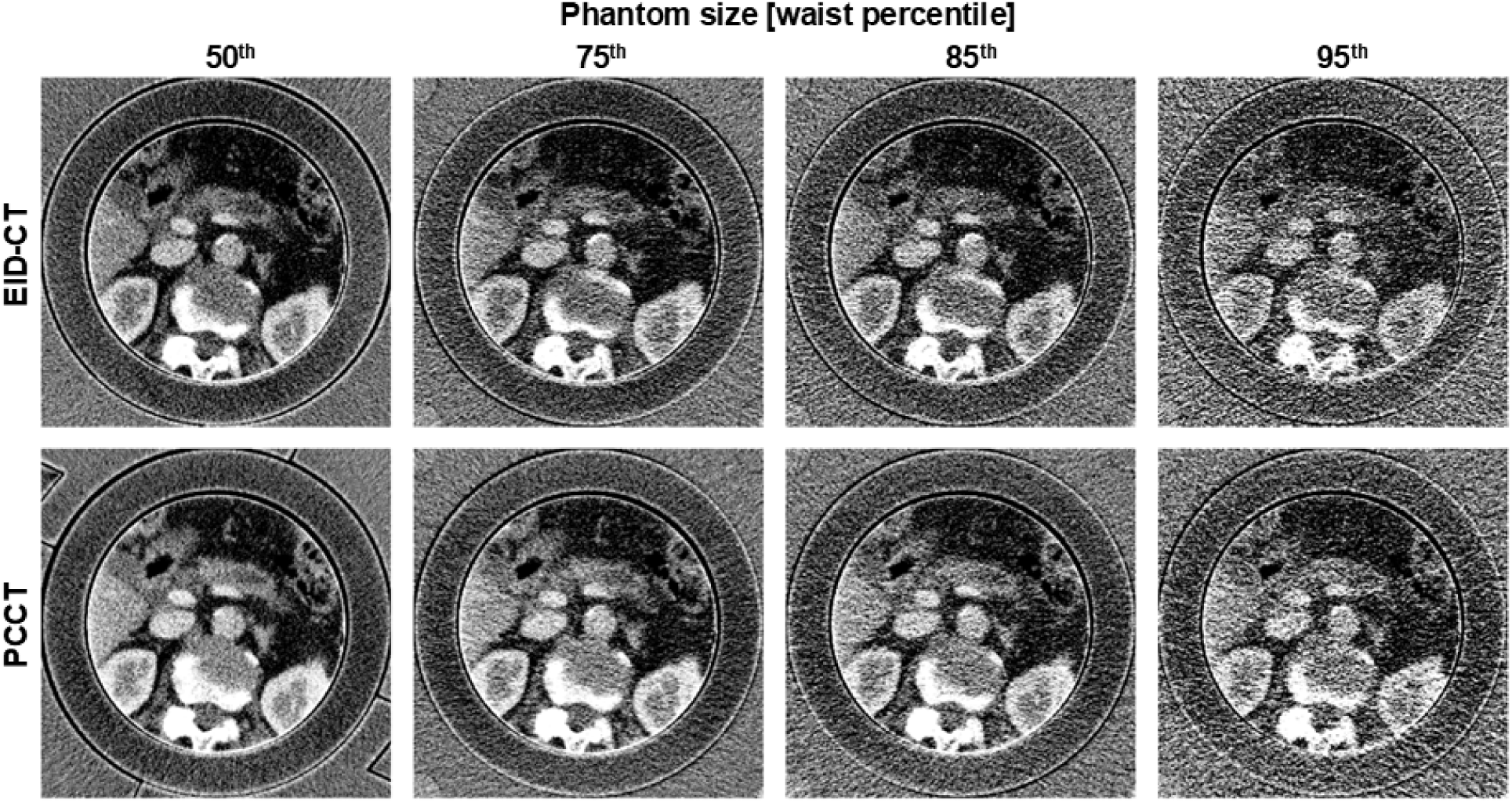
FBP images of the pancreas phantom with different phantom sizes and scanners. For images acquired at a CTDI_vol_ of 4 mGy, PCCT demonstrated reduced noise compared to EID-CT for each phantom size. WL/WW: 40/350 HU.

### Image acquisition

The different sized phantoms were scanned on both an EID-CT (Aquilion One Insight Edition, Canon Medical Systems Corporation) and a CZT-based PCCT prototype at a tube voltage of 120 kVp. Volumetric CT dose index (CTDI_vol_) values of 0.5, 1, 2, 4, 6, 10, 14.5, and 19.4 mGy were utilized. Scans were performed at each CTDI_vol_ five times to ensure adequate statistics. To determine the dose reduction between EID-CT and PCCT for each phantom, additional scans were performed on both scanners to match the reference noise on PCCT filtered back projection (FBP) images obtained at the diagnostic reference level for abdomen/pelvis CTs (16 mGy)^26^. The resulting CTDI_vol_ was then recorded to calculate the percent dose reduction for each phantom size. All images were reconstructed with FBP at a field of view of 350 mm, a slice thickness of 3 mm, and a body reconstruction kernel (FC03). Additional acquisition and reconstruction parameters can be found in Table 1.

**Table 1.** Acquisition and reconstruction parameters.

| **Scanner type** | **EID-CT** | **PCCT** |
| --- | --- | --- |
| **Scanner model** | Aquilion One Insight Edition | TSX-501R |
| **Tube voltage [kVp]** | 120 |  |
| **Spiral pitch factor** | 0.813 | 0.828 |
| **Collimation** | 80 x 0.5 mm | 192 x 0.206 mm |
| **Reconstruction algorithm** | FBP |  |
| **Field of view [mm]** | 350 |  |
| **Slice thickness [mm]** | 3 |  |
| **Matrix size** | 512 x 512 |  |
| **Pixel spacing [mm]** | 0.68 |  |

### Image analysis

To characterize the noise for each scanner, phantom size, and dose combination, the noise power spectrum^27^ (NPS) was determined using four square regions of interest (ROI) with a width of 40 px. They were placed in the background of the 20 cm phantom across ten central, non-overlapping slices and five scan repetitions (200 total ROIs). The average image was then subtracted from each ROI to isolate noise. Using these difference images, the 2D NPS was calculated and radially averaged to generate the 1D NPS and determine the peak frequency. To assess differences in noise texture, the 1D NPS was displayed for each scanner and phantom size at 4 mGy. The peak frequency for each phantom size and scanner combination was determined and averaged across phantom sizes to characterize noise textures.

ROIs were placed on both the pancreas phantom and the inserts to characterize the dose efficiency of PCCT. For the pancreas phantom, rigid registration was utilized to match image slices between scanners and phantom sizes and correct for small differences in phantom rotation. ROIs were then placed on non-tumor and tumor regions of the pancreas. Standard deviation (noise) for non-tumor and tumor regions was then measured. For the inserts, ROIs were placed automatically for each with in-house software. Standard deviation (noise) was then measured across 10 central, non-overlapping slices and averaged. Differences in noise between scanners for a given phantom size and dose were calculated to quantify the noise reduction with PCCT. A scatter plot was also utilized to compare noise across dose between scanners in both the non-tumor ROI and inserts. Error bars corresponded to the standard deviation across repetitions and across repetitions and slices for the non-tumor ROI and inserts, respectively. Additionally, noise for inserts was plotted for different phantom sizes to demonstrate the relationship between noise reduction and phantom size at different doses. Noise matched data was then utilized to determine potential dose reduction with PCCT by calculating the percent dose reduction from EID-CT to PCCT at each phantom size.

To ascertain the effect of scanner technology on non-Poisson noise at low doses (< 2 mGy) and each phantom size, a linear fit was first applied to the inverse square relationship between noise and CTDI_vol_ that characterizes Poisson noise as described in Liu et al^18^. Each linear fit incorporated noise from higher doses (CTDI_vol_ ≥ 2 mGy) as Poisson noise dominates at higher doses. At low doses (CTDI_vol_ < 2 mGy), non-Poisson system noise, such as electronic noise, is appreciable and was characterized by the root mean square error (RMSE) from the linear fit. The Shapiro-Wilk test was then performed to determine normality of the RMSE. As the data demonstrated normality (p > 0.05), a paired t-test was utilized to evaluate the effect of scanner technology on the RMSE/non-Poisson noise for each phantom size. A Bonferroni correction was applied to account for multiple comparisons such that a p-value of less than 0.0125 denoted significance.

## Results

### Noise reduction

Noise decreased with PCCT compared to EID-CT for all phantom sizes and doses, underscoring the improved dose efficiency with PCCT (Figure 3). The 1D NPS highlighted not only a difference in noise texture between EID-CT and PCCT with average peak frequencies of 0.29 ± 0.01 and 0.22 ± 0.01 mm^-1^, respectively, but also an overall decrease in noise at all frequencies with PCCT (Figure 3A). This noise reduction with PCCT was also observed in both the pancreas phantom (Figure 3B) and inserts (Figure 3C) at each dose. At 19.4 mGy, non-tumor noise for EID-CT and PCCT averaged 27 ± 1, 32 ± 2, 39 ± 2, and 59 ± 6 HU and 19 ± 2, 25 ± 1, 30 ± 2, and 41 ± 8 HU for the 50^th^, 75^th^, 85^th^, and 95^th^ percentile phantoms, respectively. Similarly, noise in inserts at 19 mGy decreased 5, 8, 12, and 23 HU from EID-CT to PCCT for the varying phantom sizes, respectively.

**Figure 3.**
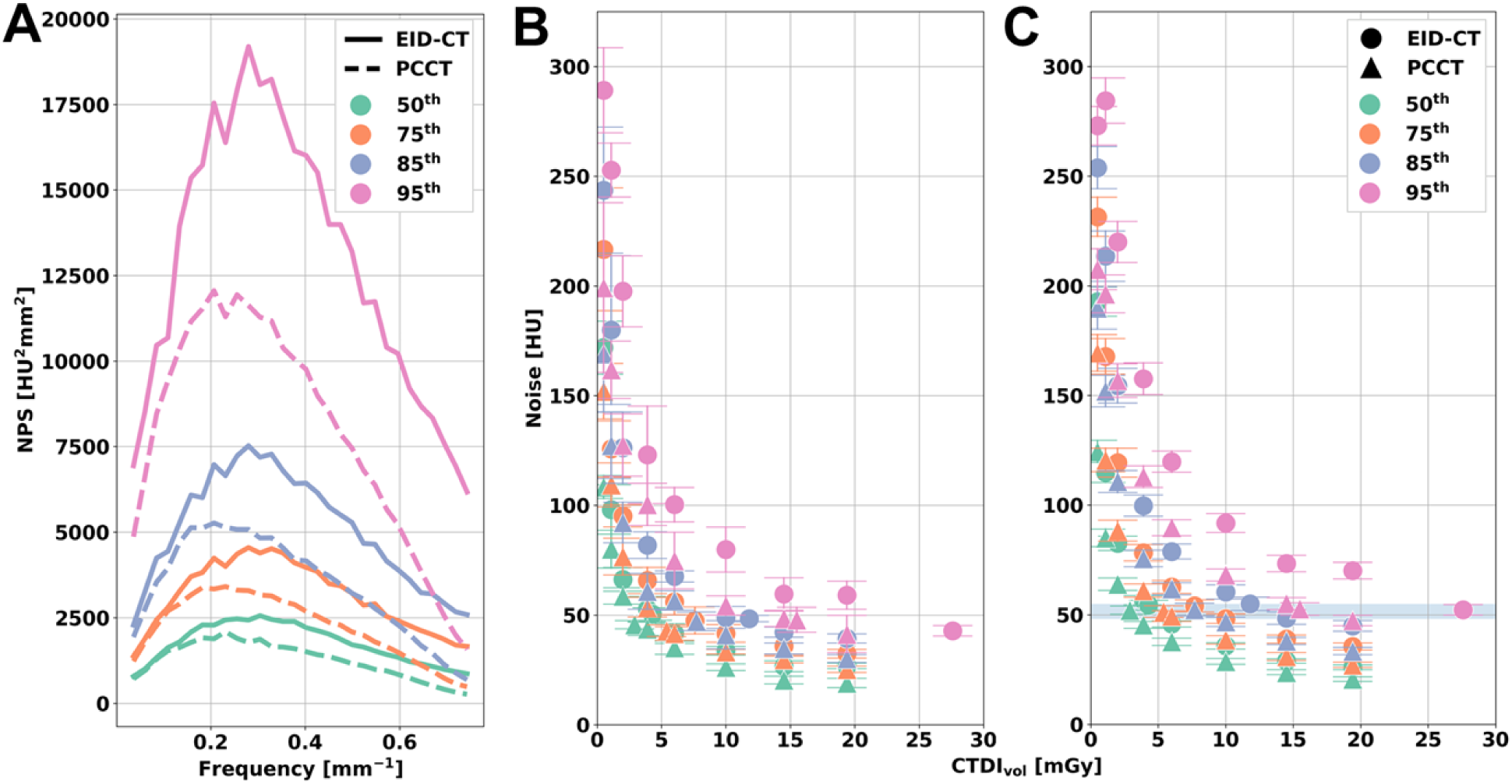
Noise-dose relationship across different scanners and phantom sizes. At each noise frequency, the noise power for PCCT was reduced compared to that of EID-CT for each phantom size (A). PCCT noise decreased compared to that of EID-CT for each phantom size and dose for both the non-tumor (B) and the brain insert ROIs (C). Error bars corresponded to the standard deviation of measured noise across repetitions (B) and repetitions and image slices (C). The blue shaded region (C) represents the noise level utilized to determine potential dose reduction with PCCT.

As dose decreased and phantom size increased, noise reduction with PCCT for both phantoms was amplified (Figure 4). Noise reduction with PCCT improved from 5 to 8, 8 to 13, 10 to 17, and 23 to 30 HU for the 50^th^, 75^th^, 85^th^, and 95^th^ percentile phantoms, respectively, as the dose decreased from 19.4 to 6 mGy. At CTDI_vol_ < 6 mGy, noise reduction for the 50^th^ percentile phantom increased to 19 and 30 HU for 2 and 1 mGy, respectively. For larger phantoms, the effect was more pronounced with noise reductions of 63 and 88 HU at 2 and 1 mGy, respectively, for the 95^th^ percentile phantom. As a result, the corresponding average percent noise reduction across doses was 22% ± 6%, 23% ± 3%, 25% ± 3%, and 28% ± 3% for 50^th^, 75^th^, 85^th^, and 95^th^ percentile phantoms, respectively.

**Figure 4.**
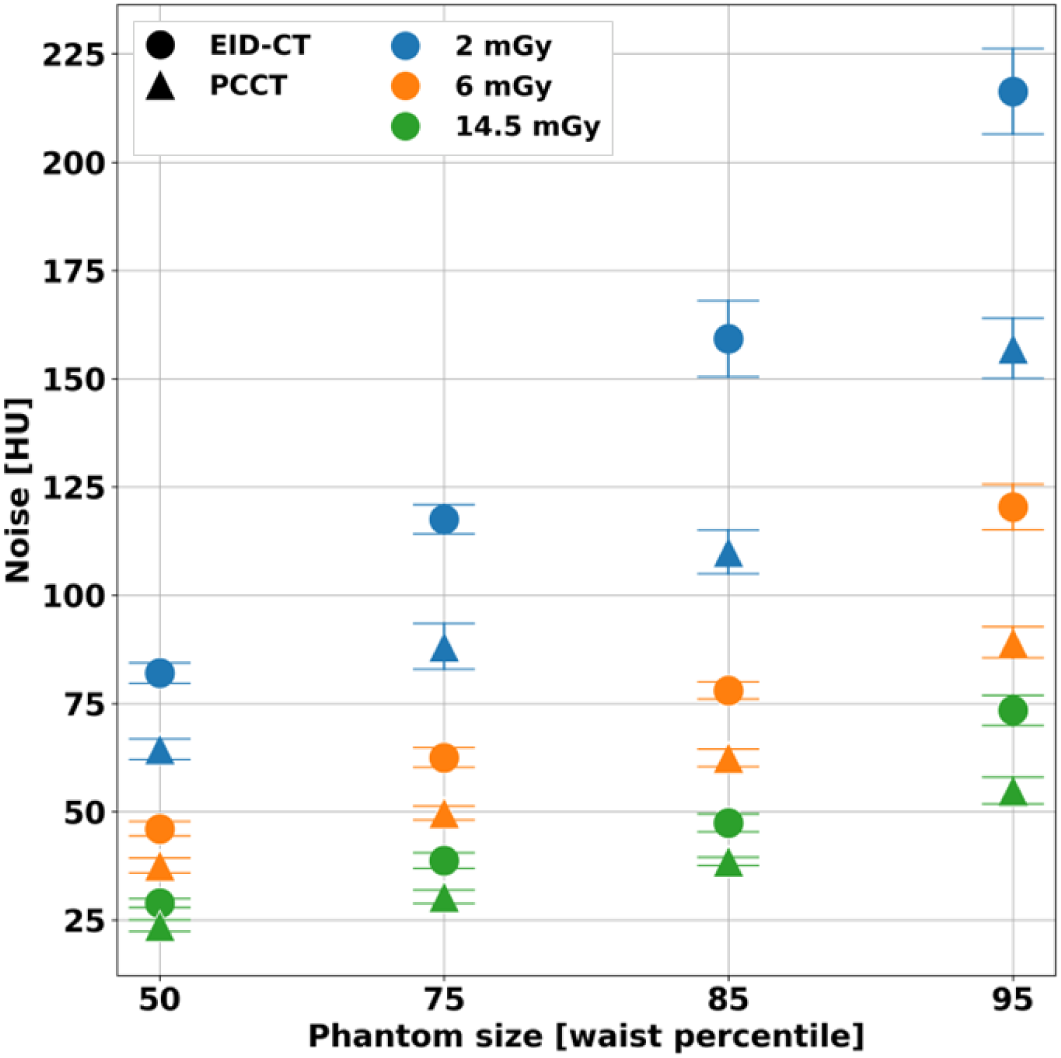
Noise for different phantom sizes. Noise in the soft tissue insert decreased for PCCT compared to EID-CT at each dose. The noise reduction with PCCT increased with enlarging phantom sizes and decreasing radiation dose, demonstrating dose efficiency and additional benefit of PCCT at larger patient sizes. Error bars corresponded to the standard deviation of measured noise across image slices and repetitions.

### Non-Poisson noise

PCCT further demonstrated improved dose efficiency with reduced influence of non-Poisson noise at low doses (Figure 5). The inverse square relationship between noise and dose for CTDI_vol_ ≥ 2 mGy was recapitulated for all inserts, phantom sizes, and scanner technologies with Pearson’s correlation coefficients greater than 0.99. At low doses (CTDI_vol_ < 2 mGy), deviations from the noise-dose relationship were less apparent with PCCT than EID-CT, indicating the reduced influence of non-Poisson noise with PCCT. With the 50^th^ and 75^th^ percentile phantoms, average RMSE across inserts significantly decreased with PCCT from 19 ± 2 to 5.0 ± 0.3 HU (p = 5 x 10^-9^) and from 15 ± 4 to 8 ± 2 HU (p = 4 x 10^-3^). As the phantom size increased, deviation from the noise-dose relationship increased for PCCT but remained significantly less than that of EID-CT. Average RMSE across inserts were 10 ± 6 (p = 6 x 10^-4^) and 43 ± 3 HU (p = 7 x 10^-12^) less with PCCT compared to EID-CT for the 85^th^ and 95^th^ percentile phantoms, respectively, exemplifying the reduction in non-Poisson noise that contributes to the enhanced dose efficiency of PCCT.

**Figure 5.**
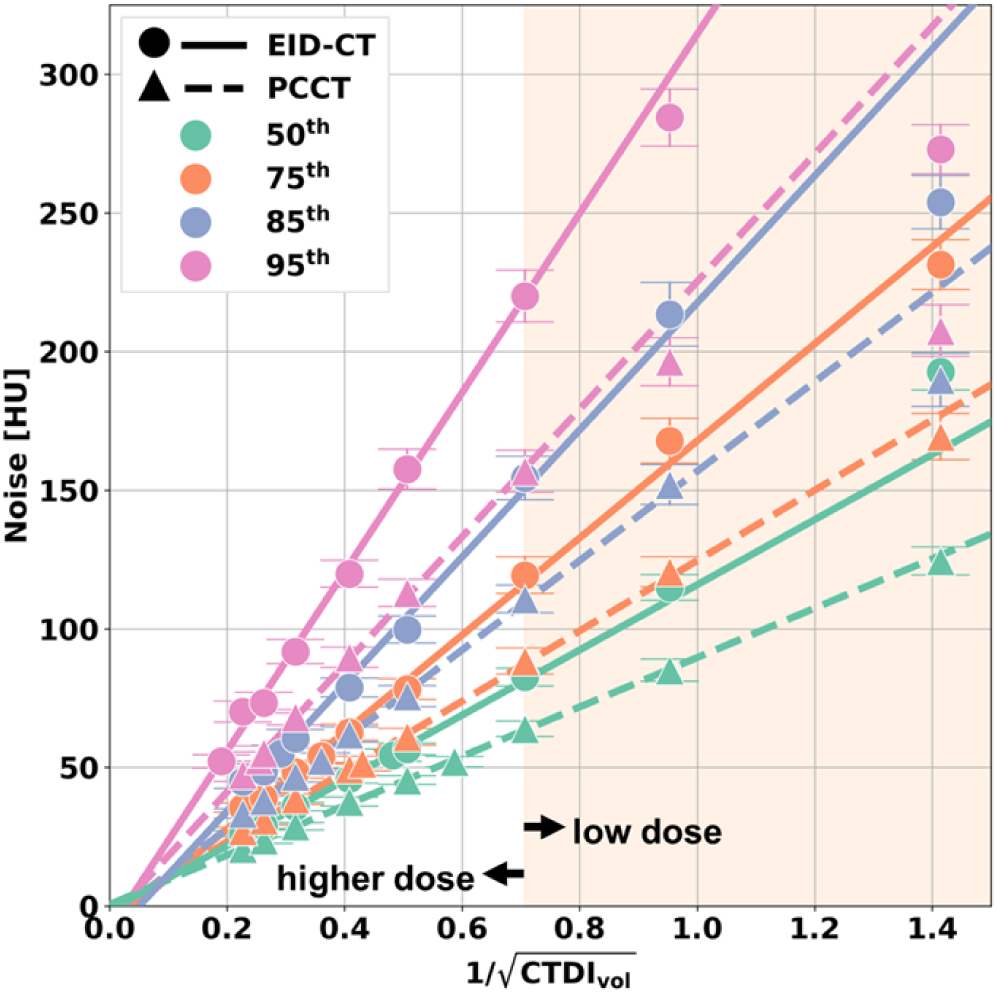
Deviations from the expected relationship between noise and dose. Dose dependence of noise measured in the brain insert was characterized by applying a linear fit for CTDI_vol_ ≥ 2 mGy (white area of the plot) for each scanner and phantom size. Noise for CTDI_vol_ < 2 mGy (low dose, orange plot area) was not included in the linear fit. At low doses, PCCT demonstrated reduced deviations from the Poisson distributed noise model compared to EID-CT.

### Dose reduction

At a matched noise level, PCCT enabled dose reduction across all phantom sizes (Figure 3C). Noise of the 95^th^ percentile phantom on PCCT FBP images at the diagnostic reference level (reference noise) measured 53 ± 3 HU. For corresponding noise-matched dose levels, noise averaged 53 ± 1 HU across phantom sizes and scanner technologies. To match the reference noise, an EID-CT dose of 27.6 mGy was required for the 95^th^ percentile phantom, corresponding to a 12.1 mGy or 44% reduction in dose with PCCT. At smaller phantom sizes, the reference noise level was met with PCCT at a CTDI_vol_ of 2.9, 5.4, and 7.7 mGy for the 50^th^, 75^th^, and 85^th^ percentile phantoms, respectively. In contrast, EID-CT doses of 4.3, 7.7, and 11.8 mGy were requiredto obtain the reference noise level. Consequently, the dose reduction decreased with phantom sizes with dose reductions of 1.4, 2.3, and 4.1 mGy for the 50^th^, 75^th^, and 85^th^ percentile phantoms, corresponding to an average dose reduction of 32% across the three phantom sizes.

## Discussion

PCCT enhanced dose efficiency given its reduced noise, including non-Poisson noise, compared to that of EID-CT across phantom sizes and radiation doses. The noise reduction with PCCT particularly improved with increasing phantom size and decreasing dose, presenting a potentially important benefit when imaging large patients. In addition, at low doses, PCCT reduced non-Poisson noise, such as electronic noise, further contributing to dose efficiency and highlighting potential advantages for low-dose applications.

Compared to previous prototype and current clinical PCCT implementations, the CZT-based PCCT prototype demonstrated similar noise reduction capabilities relative to its EID-CT counterpart. Initial phantom studies exhibited 10 – 40% noise reduction^14,16,17,19,22,28,29^. In a prototype CdTe system utilizing a 30 cm diameter phantom and FBP reconstructions, PCCT noise was consistently lower compared to that of EID-CT, resulting in noise reductions of 22% and 24% at 6 and 1.7 mGy, respectively^28^. At a comparable phantom diameter of 32 cm (50^th^ percentile phantom), this study demonstrated a similar noise reduction of 18% at 6 mGy but improved noise reduction of 23% and 26% at 2 and 1 mGy, respectively. The trend of improved noise reduction with decreased radiation dose was also observed in a first-generation commercial PCCT system with noise reductions of 30% and 38% with a 31 cm diameter phantom^16^ and 20% and 33% with a 15 cm phantom^14^ at 6 and 1.8 mGy, respectively. Noise reduction with PCCT was further replicated in a few patient studies^29,30^. Likewise, our study recapitulated the reduction in electronic noise to those highlighted in previous studies with prototype^19^ and clinical PCCT systems^14,18^. A significant decrease in RMSE, which quantifies non-Poisson noise, was observed in both a first-generation clinical PCCT^18^ and a CZT-based PCCT prototype, emphasizing the technical advantage of PCCT at low dose.

While previous studies evaluated PCCT with phantoms that correspond to an average adult size or smaller, our assessment of PCCT utilized phantom sizes that mimic larger patient habitus, highlighting additional advantages of PCCT for obese patients. Imaging of obese patients is challenging: increased attenuation often requires higher radiation doses in order to achieve diagnostic image quality^6^. However, the dose required to achieve diagnostic image quality may not be attainable with current EID-CT technology, leading to poor image quality and artifacts^7^. PCCT, on the other hand, has demonstrated enhanced dose efficiency that improves noise and image quality at a given dose compared to EID-CT. Our work especially underscores qualitative and quantitative improvement of noise with larger phantoms, enabling an average potential dose reduction of 35% without compromising image quality. Comparable dose reduction ranging from 32% to 39% has been reported for abdominal imaging^16,17,20,21^ while lung imaging has demonstrated higher potential dose reductions up to 66%^22,31,32^. This dose and noise reduction underline the improved dose efficiency that may lead to better visualization of anatomical structures critical for accurate diagnosis and disease monitoring for both the imaging of obese patients and low-dose applications. In addition, noise reduction with PCCT may enable high resolution imaging in obese patients, which was previously not achievable with EID-CT.

This study has a few limitations. First, this study only presents an evaluation of noise in conventional images available with the current CZT-based PCCT prototype. Future evaluations will incorporate spectral results to capture the advantages of PCCT more fully on clinically relevant image types. These spectral results, particularly virtual monoenergetic images, also reduce beam hardening, which may improve image quality for obese patients. Second, only FBP reconstructions were utilized in this study to evaluate image noise and dose efficiency. Advanced reconstruction methods such as deep learning reconstruction were not included but will be evaluated in future studies to assess the combined effects of reconstruction method and detector technology on image quality and dose efficiency. Third, the adult waist circumference in the United States population was utilized to generate 3D-printed extension rings. Unfortunately, there is very little data on the distribution of water equivalent diameters of adult abdomens, BMIs, and their correlation, preventing a more accurate depiction of adult patient sizes at each percentile. Regardless, the phantom sizes in this study correspond to obese patient sizes seen in our clinical practice. Additionally, our study utilized both a geometric phantom and anthropomorphic *PixelPrint* phantom as a surrogate for patients. While these phantoms may serve as an intermediate step, findings will ultimately need to be confirmed in clinical routine. Finally, only one scan protocol and EID-CT were evaluated in this study. Future studies will expand the parameter space to better understand the effects of key acquisition and reconstruction parameters.

## Conclusion

The dose efficiency of PCCT was superior to that of EID-CT across a wide range of phantom sizes and radiation dose levels. PCCT, with enhanced dose efficiency, promotes both noise and dose reduction and may be a useful tool for the imaging of obese patients and low-dose clinical applications.

## Data Availability

All data produced in the present study are available upon reasonable request to the authors.

## Acknowledgement

We acknowledge support through the National Institutes of Health (R01EB035092) and Canon Medical Systems Corporation (Otawara, Japan).

## Abbreviations

PCCT: photon-counting CT
EID-CT: energy-integrating detector CT
CZT: cadmium zinc telluride
CTDI_vol_: volumetric CT dose index
FBP: filtered back projection
NPS: noise power spectrum
ROI: region of interest
RMSE: root mean square error

